# Cost-effectiveness of a whole-area testing pilot of asymptomatic SARS-CoV-2 infections with lateral flow devices: A modelling and economic analysis study

**DOI:** 10.1101/2021.05.10.21256816

**Authors:** Mark Drakesmith, Brendan Collins, Angela Jones, Kelechi Nnoaham, Daniel Thomas

**Author notes:** Correspondence: Mark Drakesmith < >.

## Abstract

**Background:** Mass community testing for SARS-CoV-2 by lateral flow devices (LFDs) aims to reduce prevalence in the community. However its effectiveness as a public heath intervention is disputed.

**Method:** Data from a mass testing pilot in the Borough of Merthyr Tydfil in late 2020 was used to model cases, hospitalisations, ICU admissions and deaths prevented. Further economic analysis with a healthcare perspective assessed cost-effectiveness in terms of healthcare costs avoided and QALYs gained.

**Results:** An initial conservative estimate of 360 (95% CI: 311-418) cases were prevented by the mass testing, representing a would-be reduction of 11% of all cases diagnosed in Merthyr Tydfil residents during the same period. Modelling healthcare burden estimates that 24 (16 - 36) hospitalizations, 5 (3-6) ICU admissions and 15 (11-20) deaths were prevented, representing 6.37%, 11.1% and 8.2%, respectively of the actual counts during the same period. A less conservative, best-case scenario predicts 2333 (1764-3115) cases prevented, representing 80% reduction in would-be cases. Cost effectiveness analysis indicates 108 (80-143) QALYs gained, an incremental cost ratio of £2,143 (£860-£4,175) per QALY gained and net monetary benefit of £6.2m (£4.5m-£8.4m). In the best-case scenario, this increases to £15.9m (£12.3m-£20.5m).

**Conclusions:** A non-negligible number of cases, hospitalisations and deaths were prevented by the mass testing pilot. Considering QALYs gained and healthcare costs avoided, the pilot was cost-effective. These findings suggest mass testing with LFDs in areas of high prevalence (>2%) is likely to provide significant public health benefit. It is not yet clear whether similar benefits will be obtained in low prevalence settings or with vaccination rollout.

## Introduction

The aim of mass community SARS-CoV-2 (COVID-19) testing by lateral flow device (LFD) tests is to provide a significant and sustainable reduction in the prevalence of infection in an area. By offering testing to the whole community, it is possible to identify asymptomatic infections that would not otherwise be detected. Through prompt isolation and contact tracing of these newly identified asymptomatic or pre-symptomatic individuals, community transmission may be reduced. The LFD test is advantageous because of it’s speed and ease of deployment to large populations, compared to the polymerase chain reaction (PCR) test. LFD testing is deemed particularly beneficial for groups that are more likely to be asymptomatic, such as children and students, or for key workers who are in regular contact with vulnerable individuals. Community-wide asymptomatic testing was subsequently rolled out across the UK.^1^

Prior to national roll out, pilots were conducted in Liverpool^2^ and later in the Merthyr Tydfil and the Lower Cynon Valley area of South Wales during November to December 2020..^3^ Merthyr Tydfil had, at the time, one of the highest incidence rates of COVID-19 in the UK along with associated illness and deaths (248.6 cases per 100,000 in the first week of the pilot^4^). The Welsh Index of Multiple Deprivation shows that Merthyr Tydfil has some of the most deprived areas in Wales.^5^ In addition, smoking rates are high in both areas in comparison to the health board and the Welsh average, and there are high levels of overweight and obesity and higher prevalence of long term conditions, all factors associated with poor COVID-19 outcomes.^5^

The LFD test has been criticised for its poor sensitivity. Initial reports from the Liverpool pilot reported the true positive rate of LFD tests was 99.9%, but a false negative rate of 48.9%.^6^ A study of LFD tests in UK university students found sensitivity can be as low as 3%.^7^ The sensitivity of the test is highly dependent on the expertise of who the test is administered by.^8^ LFD tests sensitivity is significantly influenced by viral load.^7^ Viral load peaks in asymptomatic cases are lower than for symptomatic cases, affecting the usefulness of LFDs as a screening tool. Furthermore, the false sense of reassurance a negative test result could provide may lead to more risky behaviours and reduced adherence to social distancing and lockdown restrictions. Clear information regarding the low sensitivity of LFD tests is not always readily available to the public.^9^ Consequently, the use of LFD mass testing as a screening tool has provoked controversy in the medical community and the media.^10^ It has been argued that mass testing with LFD devices is not cost effective, with one estimate of the cost of detecting a single asymptomatic case by LFD mass testing as much as £30,000.^12^ However, such estimates do not take into account the potential costs saved in terms healthcare burden and quality life years lost though onward transmission that would have occurred had mass testing not identified these cases.

There has been previous studies that have evaluated the merits of the LFD testing in terms of sensitivity and specificity,^6^ but not on their cost effectiveness in the context of mass testing. Most existing studies assessing cost-effectiveness of mass testing focus on screening exercises with PCR (see 13 for a review). One recent modelling study by 14 assessed the cost-effectiveness of LFD testing in a range of hypothetical scenarios in the USA. Their results also suggest mass testing by LFD is highly cost-effective, with repeated testing over a 28-day increment with 2-week isolation for positive cases suggested as the most cost-effective. A cost-benefit analysis of mass testing in Barcelona, suggests that absolute cost-benefit ratio of asymptomatic mass testing was low (<1), but when including monetised healthcare outcomes in the analysis, showing a high (>1) cost-benefit ratio.^15^ A comparable economic analysis has not yet been conducted on the Liverpool pilot, however an interim analysis using Bayesian time-series modelling estimated a small non-significant reduction in cases and hospitalisations due to mass-testing.^2^ However, as illustrated by the Barcelona study, by not considering monetised healthcare outcomes, it is difficult to infer whether economic impacts were significant.

In the present study, we assess the impact of the whole area testing pilot in Merthyr Tydfil by modelling the number of onward infections prevented using the reproduction number (Rt) in Merthyr Tydfil at the time and applying a series of assumptions on the natural history of the infection and the performance of the test. These estimates are then applied to a time-lagged regression model to estimate the number of hospitalizations, ICU admissions and deaths that would have arisen from the prevented cases. We also perform sensitivity analysis on some parameters that underlie the assumptions made in the analysis and derived estimates from likely worst-case and best-case scenarios. Finally, the cost effectiveness, in terms of incremental cost effectiveness ratio (ICER) per QALY gained is estimated.

## Method

### Study design

Modelling and economic analysis, using original data collected as part of a mass testing pilot.

### Ethics, Data privacy and information governance

The study was reviewed by the Public Health Wales Research and Development Office and determined to be usual practice in public health, and therefore did not require external NHS ethics committee approval. Investigation of communicable disease outbreaks and surveillance of notifiable disease is permitted under Public Health Wales Establishment Order. Data were held and processed under Public Health Wales information governance arrangements in compliance with the Data Protection Act, Caldicott Principles and Public Health Wales guidance on the release of small numbers. No data identifying protected characteristics of an individual were released outside Public Health Wales.

### Sample data

Lateral flow device (LFD) tests, using the INNOVA SARS-CoV-2 Rapid Antigen test device, were performed as part of the whole area testing pilot that took place between 21st November 2020 and 20th December 2020 in Merthyr Tydfil and lower Cynon Valley. Testing was offered to individuals aged 11 or over, living or working in the area and who were not experiencing symptoms of COVID infection at the time of testing. Full details of the pilot are provided in the pilot evaluation.^3^ For ease, the present analysis was restricted to Merthyr Tydfil only as this is a whole self-contained local authority, whereas lower Cynon Valley comprises a sub-region of the Rhondda Cynon Taf local authority. Of 48,834 tests that took place during the pilot, 33,822 were taken at one of 14 test centres in Merthyr Tydfil. Of these, 712 positive LFD tests who reported being asymptomatic were used for the analysis. This represents 2.1% of tests performed.

### Analysis

#### Estimates of cases prevented

All analysis was carried out using R software. Assumptions made in estimating cases prevented are given in Box 1. LFD positive asymptomatic cases during the testing period were counted for each day. From this, the number of LFD positive tests that were confirmed by a PCR test, either on-site or by an independent test no more than 14 days later, were counted. From this number, the number of true positives were estimated. Firstly we identified all LFD test positives with a confirmatory PCR test result (classified as true positives). Secondly, we estimated the number of true positives in those who did not have a confirmatory PCR test from the positive predictive value (PPV) of the LFD test (0.98). This was estimated from the proportion of LFD positives that had follow-up PCR (539), that were confirmed true positives (528).

##### Box 1: Assumptions of the analysis

A summary of the assumptions underlying the model is provided below.

1. All infected persons are equality infectious irrespective of age, gender, demographic variables, etc.
2. Infectability of asymptomatic cases are 58% of symptomatic cases. ^17^
3. The number of infections created by an infectious person is determined by the Rt value on the day of their LFD positive test result.
4. The daily count of new infections arising from infectious persons is a fixed Log Normal distribution with a median of 5 days and up to 14 days long. ^19^
5. An infectious person became infectious 5 days before the LFD positive test result and up to 9 days after (14 days in total). Only infections taking place after the LFD positive test are considered preventable.
6. All infected persons not exhibiting symptoms will not self-isolate, therefore will contribute to subsequent infections.
7. Most persons exhibiting symptoms will self-isolate, but some will still contribute to secondary transmission (each symptomatic case will contribute 0.2 secondary cases in contacts).
8. The proportion of positive cases that will develop symptoms remains constant (0.44). ^16^
9. All cases reported asymptomatic are truly asymptomatic.
10. All positive LFD tests with confirmatory PCR test are true positives.
11. Of the positive LFD tests without confirmatory PCR test, a fixed proportion will be true positives (0.98)

A proportion of asymptomatic cases will go on to develop COVID-19 symptoms. The number expected to remain asymptomatic was estimated by scaling the number of true positives by the proportion of asymptomatic cases that did not report having symptoms when interviewed coincidentally as part of a case-control study that took place during the testing pilot. From 198 LFD positive results, 87 (44%) reported symptoms when followed up.^16^ Asymptomatic infections are assumed, had mass testing not taken place, to not self-isolate and therefore would contribute fully to the next generation of infections. A small proportion of secondary infections (0.2) were assumed for symptomatic cases, resulting from infectious contact made prior to onset of symptoms or as a failure of self-isolation. The sum of these two count constitutes the initial set of cases identified though the testing pilot but not directly prevented (also referred to as 0th generation infections). The process for obtaining this count is visualised in the flowchart in Figure 1a.

**Figure 1:**
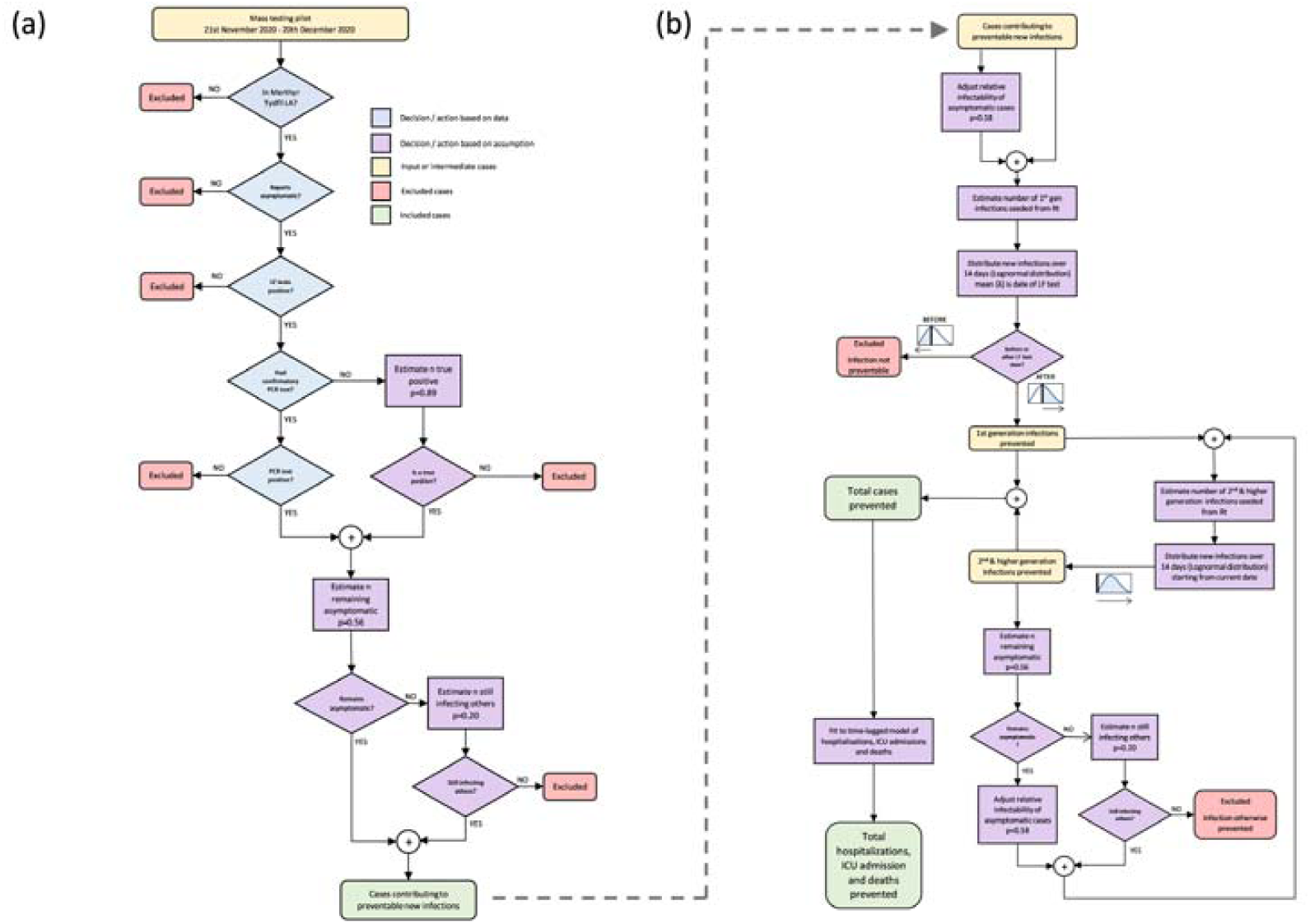
Flowchart of process for (a) estimating number of cases detected by mass testing that would not otherwise be detected and for (b) estimating number of cases prevented, and other healthcare outcomes. This process is followed iteratively for each day in the testing period.

From the initial set of cases, the 1st generation infections that would have occurred from these cases is estimated by multiplying by the reproduction number (Rt - mean and 95% CI) for that day by the number true positive LFD test cases. A scaling factor of 0.58 is also applied to take into account the relative reduction of infectability of asymptomatic cases compared to symptomatic cases.^17^ The daily reproduction number Rt was estimated using the “EpiEstim” R package^18^ based on actual PCR-tested case counts in Merthyr Tydfil local authority area (see Figure 2a).

**Figure 2:**
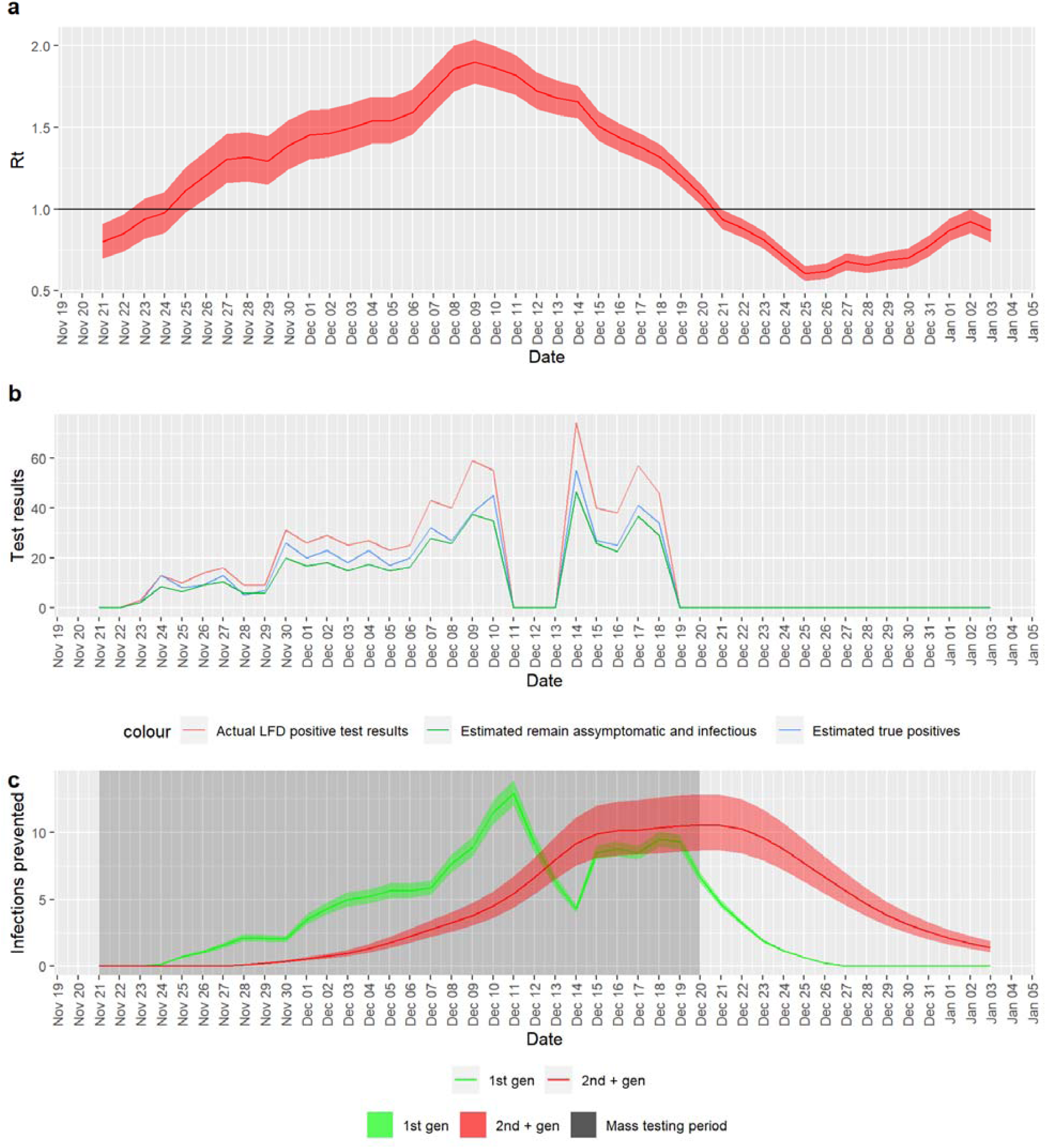
(a) Estimated Rt in Merthyr Tydfil over the testing period. (b) Positive lateral flow test results from community testing pilot in Merthyr Tydfil, with estimates for true positives and proportion remaining asymptomatic. Note that missing data in the weekend of 11th to 13th December is due to temporary suspension of the mass testing on these dates. (c) Daily estimates of 1st generation and 2nd + higher generation infections prevented

The number of subsequent (2nd and higher generation) infections were estimated by iteratively multiplying the estimated Rt by the estimated number of prevented infections from the previous day, each time scaling the number of cases from the previous generation by the proportions of asymptomatic and symptomatic-infectious cases, and scaling the asymptomatic portion by the relative infectability of asymptomatic cases, as done for the 0th generation.

The number of infections estimated prevented for a given day is the sum of the 1st generation and 2nd and higher generation infections. 0th generation infections are excluded from this sum as they occurred before the LFD test and therefore cannot be considered preventable.

The number of infections estimated for a given day is then distributed over an infection window of 14 days, which has a log-normal distribution (*µ*=1.63, *σ*=0.5), with a median of 5 days. The parameters for this distribution were taken from the meta-analysis of McAloon et al ^19^. For 1st generation infections (infections arising direct from the LFD positive cases) the infectious period is assumed to be centred around the LFD test date, to take into account the likely delay between the date of infection and the date of the LFD test. Only the infectious period after the LFD test date is considered as any new infections generated before the LFD test would not have been prevented. For 2nd and higher generation infections, the whole infection window is considered and the start of the infection window is the date the infection is seeded. A flowchart depicting this process is shown in Figure 1b and a mathematical description is given in the appendix.

The proportion of symptomatic cases that contribute to new infections was estimated from contact tracing data (restricted to the period of the mass testing exercise and to the Merthyr Tydfil local authority area). The contacts for all cases that reported symptoms were cross referenced to identify those that also have a PCR-confirmed positive test result. The proportion of contacts of symptomatic cases that were themselves identified had a positive test results was 0.2.

Some of the parameters underlying the assumptions of the analysis have a high degree of uncertainty. Three parameters were highlighted for sensitivity analysis and used to produce worst-case and best-case scenarios for the number of cases prevented (see Appendix B for details). These additional scenarios were included in the subsequent analyses.

#### Estimates of healthcare burdens prevented

The estimated number of cases prevented were further used to predict three healthcare outcomes: Hospitalisations, ICU admissions and deaths. A time-lagged log-linear regression model was trained on corresponding surveillance data for residents of Merthyr Tydfil Local Authority.^4^ Separate models were built for each healthcare outcome with multiple lagged case counts treated as the predictor variables and the burden measure as the outcome measure. Hospitalisation data were restricted to those where a positive PCR test was obtained no later than 2 days after admission. This rules out likely nosocomial infections, as opposed to those that are community-acquired. All data were log transformed and smoothed with a LOESS filter (span = 0.2). Models are built from time-lagged case counts with delays of 12-50, 24-50 and 30-50 days for hospitalizations, ICU admissions and death, respectively. These lag ranges were determined to be optimal by testing multiple lag windows of 1-50 days and comparing the AIC/BIC values.

The estimated number of cases prevented computed in the previous section were then fed into the model to predict daily counts of each healthcare burden prevented. The counts are then summed across all days modelled to obtain a final count of each healthcare burden.

#### Economic analysis

Standard cost effectiveness analysis (sometimes called cost utility analysis) methods were used.^20^ There have been several attempts to attach costs to COVID-19 outcomes.^21^ For this work, we used a ‘bottom up’ method using Wales-specific medical costs and intervention costs collected as part of the mass testing pilot. The cases and associate hospital burdens were multiplied by estimated costs and QALYs lost. This was done for the main estimate and the best- and worst-case scenarios derived from the previous sensitivity analysis. The QALY losses from COVID-19 cases included cases, deaths, hospital admissions and a conservative (low) estimate of the lost QALYs from long COVID-19. The programme costs for the mass testing pilot in Merthyr Tydfil were estimated as £515,688.^3^ Costs were measured in GBP, at 2020 prices. QALYs lost from COVID-19 deaths were discounted at 1.5% per annum in line with UK Treasury Green Book. Other costs and QALYs were not discounted as they were assumed to occur within-year. Table 3 shows the estimated outcomes and unit costs and QALYs for each outcome. No costs were assumed for cases that do not end in hospital or dying. Further details of the cost and QALY estimates are provided below.

#### COVID-19 community cases

COVID-19 community cases cause a 0.000889 QALY loss, equivalent to asthma for 7 days.^22^ With the model results, QALY losses for long COVID-19 are also applied to a percentage of cases.

#### Post-viral syndromes

We used 0.15 QALYs lost (0.3 utility score decrement for six months) which is the equivalent of moderate fibromyalgia^23^ for six months,^23^ but also may be similar to other syndromes that are similar to ‘long COVID-19’ post-viral syndromes. We had clinical input into using fibromyalgia: as a similar syndrome but in reality long COVID-19 may be several distinct syndromes. We applied this to 10% of COVID-19 cases as ONS data suggests that around 10% of people in the COVID-19 infection survey report symptoms for 12 weeks or more.^24^

#### Deaths

Deaths result in a QALY loss of 7.24 QALYs per death, based on data from the Secure Anonymised Information Linkage (SAIL) databank that estimated mean years of life lost of 9.97 when compared with age- and sex-specific life tables, multiplied by average health-related quality of life (HRQoL) utility index for people aged 75 and over in the UK (0.726).^25^ This may be a high estimate as it includes age but does not directly factor in co-morbidities in terms of mortality risk. Discounted at 1.5% per annum this is 6.78 QALYs. COVID-19 deaths results in an excess healthcare cost of £232.^26^ This is likely a low estimate.

#### Hospital and ICU admissions

We assume 0.01126 QALYs lost per admission and 0.274 HRQoL loss for 15 days, based on people in hospital for pneumonia.^27^ Cost per admission of £7,085 is (NHS Wales Finance Delivery Unit). We assume 1.402 HRQoL loss for 9 days^27^ (Note this means that the QALYs for this health state are ‘worse than death’). Cost per admission is £22,198 (NHS Wales Finance Delivery Unit). Our hospital costs are higher than other estimates like NHS tariff/ reference costs as they include costs of PPE and other costs associated with the pandemic.

## Results

### Estimates of cases prevented

A total of 33,823 lateral flow device (LFD) tests were performed in Merthyr Tydfil County Borough between 21st November 2020 and 20th December 2020, of which 712 were positive and reported asymptomatic. Of these 697.4693878 were estimated to be true positives (528 confirmed by PCR and an additional 169 estimated from PPV). Of these, 391 were estimated to not subsequently develop symptoms and contribute to secondary infections. In addition, an estimated 62 of those developing symptoms would still contribute to secondary infections. Figure 2b shows the daily count of these numbers over the testing period. Summary counts are provided in Table 1.

**Table 1:** Estimated cases prevented.

| Description | n | upper CI | lower CI |
| --- | --- | --- | --- |
| Actual case count | 2900 |  |  |
| LFD test performed during mass testing | 48834 |  |  |
| - of which were in the Merthyr Tydfil LA | 33823 |  |  |
| - of which were asymptomatic and tested positive | 712 |  |  |
| - of which had a confirmatory PCR test result | 528 |  |  |
| - of which had no confirmatory PCR test | 173 |  |  |
| - of which were estimated to be true positives from PPV | 169 |  |  |
| - of which are true positives (PCR-confirmed + estimated from PPV) | 697 |  |  |
| - of which were estimated to remain asymptomatic | 391 |  |  |
| - of which were estimated to become symptomatic | 306 |  |  |
| - of which were estimated to still contribute to infections | 62 |  |  |
| Infectious cases identified not otherwise identifiable. | 453 |  |  |
| 1st generation infections prevented | 169 | 182 | 156 |
| 2nd and higher generation infections prevented | 192 | 236 | 156 |
| Total estimated number of prevented cases | 360 | 418 | 311 |

Including 1st and higher generation infections, a total of 360 (95% CI: 311 - 418) infections are estimated to have been prevented in the period from 21st November 2020 to 3rd January 2021. This represents 12.4% (10.7%, 14.4%) of the actual count of clinical cases diagnosed in the same period (2,900), an effective reduction of 11% from the would-be case count without mass testing (3,260). This also represents a reduction in mean daily incidence rate of 13.9 (12 - 16.1) cases per 100,000 per day (actual mean daily incidence rate was 112 cases per 100,000 per day). This is the sum of 169 (156 - 182) estimated 1st generation infections and 192 (156 - 236) estimated 2nd and higher generation infections (see Table 1). Figure 2c shows daily estimates of infections prevented over the course of the mass testing and the following 14 days.

Table 2 shows the estimated cases prevented for worst- and best-case scenarios, the worst-case scenario being 160 (142, 180) cases prevented, which is 44% of the original estimate and 6% of the actual case count. The best-case scenario is 2333 (1764, 3115), is 648% of the original estimate and 80% of the actual case count.

**Table 2:** Total estimated cases prevented and associated healthcare outcomes for original, worst-case and best-case scenarios.

| Scenario | Cases |  |  | Hospitalizations |  |  | ICU admissions |  |  | Deaths |  |  |
| --- | --- | --- | --- | --- | --- | --- | --- | --- | --- | --- | --- | --- |
|  | n | Lower | Upper | n | Lower | Upper | n | Lower | Upper | n | Lower | Upper |
| Original | 360 | 311 | 418 | 24 | 16 | 36 | 5 | 3 | 6 | 15 | 11 | 20 |
| Worst-case | 160 | 142 | 180 | 13 | 8 | 20 | 3 | 2 | 4 | 9 | 7 | 12 |
| Best-case | 2328 | 1761 | 3107 | 66 | 49 | 92 | 9 | 7 | 12 | 33 | 26 | 42 |

### Estimates of healthcare burdens prevented

The model estimates, from the cases prevented during the mass testing, a total of 24 (16 - 36) hospitalizations, 5 (3 - 6) ICU admissions and 15 (11 - 20) deaths were prevented in the period between 21st November 2020 and 10th February 2021. These represent reductions of 6.37%, 11.1% and 8.2% of the actual hospitalization, ICU admissions and deaths, respectively. The estimated time course of these three outcomes are shown in Figure 3.

**Figure 3:**
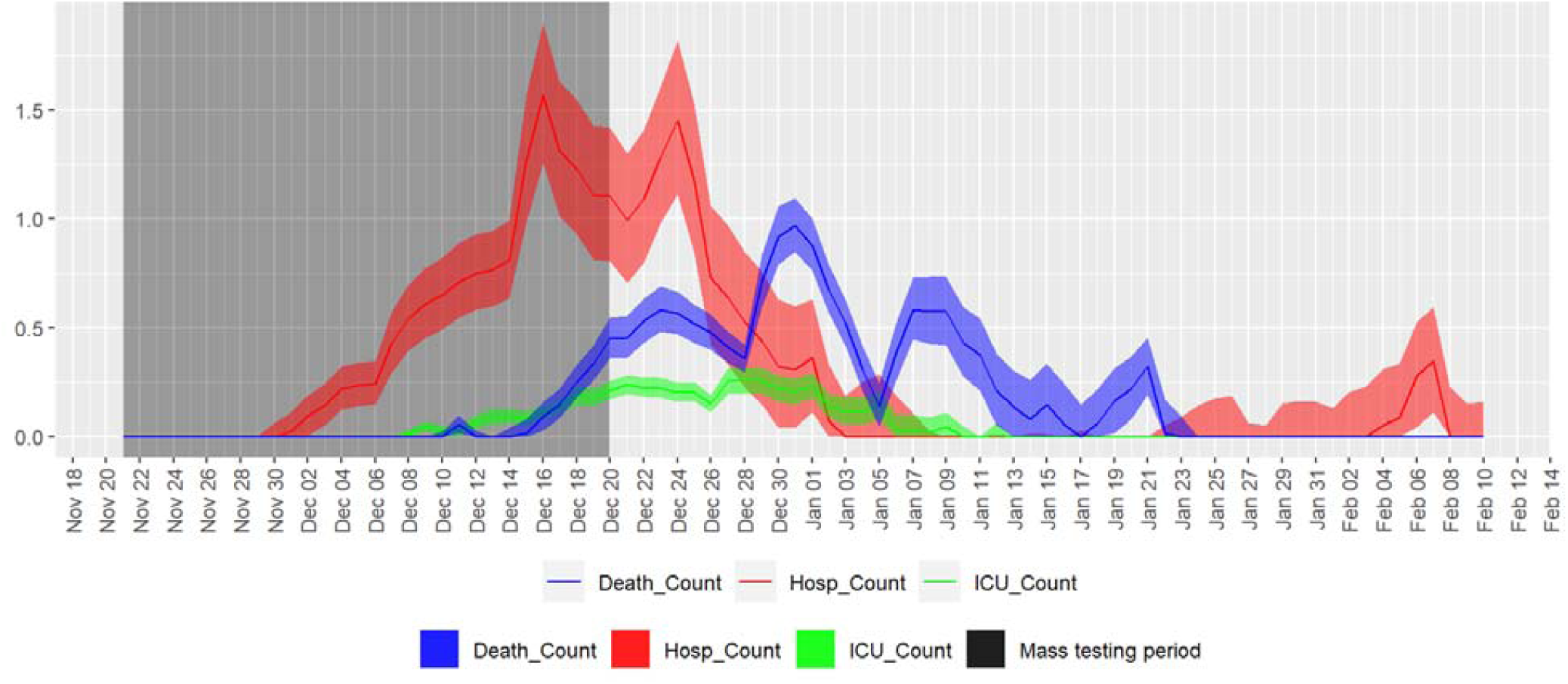
Estimates of hospitalizations, ICU admissions and deaths prevented from cases prevented by mass testing

Table 2 shows the estimated hospital burdens prevented for worst- and best-case scenarios. Healthcare burdens prevented for the best-case scenario are 66 (49 - 92) hospitalizations, 9 (7 - 12) ICU admissions and 33 (26 - 42) deaths. These represent reductions of 17.6%, 22% and 18.4% of the actual hospitalization, ICU admissions and deaths, respectively.

### Economic analysis

The programme was highly likely to be cost effective, with an incremental cost effectiveness ratio of £2,143 per QALY gained (lower confidence interval: £860, upper confidence interval: £4,175). Net monetary benefit for the intervention, which is cost savings plus the value of QALYs gained (valuing discounted QALYs at UK Treasury £60,000^28^) was £6.2million (£4.5m – £8.4m). See Table 4.

**Table 3:** QALYs and monetary costs per unit.

| Outcome | QALYs lost per case | Costs per case (£) |
| --- | --- | --- |
| Cases | 0.0159 | 0 |
| Hospitalisations | 0.0113 | 7,085 |
| ICU admissions | 0.0346 | 22,198 |
| Deaths | 6.7800 | 232 |

**Table 4:** Estimated QALYs gained, cost savings, incremental cost effectiveness ratios (ICERs) and net monetary benefit of mass testing programme in Merthyr Tydfil, for mid, high and low estimate of effectiveness.

|  | mid | low | high |
| --- | --- | --- | --- |
| QALYs gained | 108 | 80 | 143 |
| Net costs (£) | 231,188 | 333,188 | 122,814 |
| ICER (Incremental cost effectiveness ratio) cost per QALY gained (£) | 2,143 | 4,175 | 860 |
| Net monetary benefit (valuing QALYs at £60,000) (£) | 6,240,594 | 4,455,127 | 8,448,442 |
| **Sensitivity analysis** |  |  |  |
| Net monetary benefit - worst case model parameters (£) | 3,473,831 | 2,579,534 | 4,792,587 |
| Net monetary benefit - best case model parameters (£) | 15,866,328 | 12,296,102 | 20,546,837 |
| Net monetary benefit with lower QALY loss for COVID deaths (£) | 4,638,594 | 3,280,327 | 6,312,442 |
| Net monetary benefit valuing QALYs at £30,000 instead of £60,000 (£) | 3,004,703 | 2,060,970 | 4,162,814 |
| Net monetary benefit (3.5% discount rate instead of 1.5%) | 5,745,594 | 4,092,127 | 7,788,442 |

The majority of benefits (around 90%) are through the discounted QALYs lost and costs of people dying from COVID-19 (see Table 5). As a sensitivity analysis, we varied the QALYs lost from early deaths down from 6.78 down to 5 QALYs, but the net monetary benefits were still large at £4.6million. We also had a sensitivity analysis where we varied the value of QALYs down from £60,000 to £30,000 but the net monetary benefit was still positive at £3.0million. Using a 3.5% discount rate for health benefits (as recommended by NICE) instead of UK Treasury 1.5% slightly decreased the net monetary benefit from £6.2million to £5.7million.

**Table 5:** QALYs gained and cost savings for mid, low and high scenarios of mass testing intervention in Merthyr Tydfil. Negative cost savings indicate net costs.

|  | QALYs gained |  |  | Cost savings (£) |  |  |
| --- | --- | --- | --- | --- | --- | --- |
|  | mid | low | high | mid | low | high |
| Cases | 6 | 5 | 7 | - | - | - |
| Hospital cases | 0 | 0 | 0 | 170,032 | 113,354 | 255,047 |
| ICU cases | 0 | 0 | 0 | 110,989 | 66,593 | 133,187 |
| Deaths | 102 | 75 | 136 | 3,480 | 2,552 | 4,640 |
| Programme costs |  |  |  | -515,688 | -515,688 | -515,688 |
| Total | 108 | 80 | 143 | -231,188 | -333,188 | -122,814 |

## Discussion

As a result of the whole area testing pilot in Merthyr Tydfil, a non-negligible number of cases, hospitalisations and deaths, that would have otherwise occurred, were likely to have been prevented. Over one tenth of cases that would have occurred over a 6 week period were prevented. This forecast translates into a predicted reduction of 6-12% in burden on the healthcare system relative to actual healthcare burden at the time. These results were obtained for a conservative, but probable scenario. In a less-conservative, best-case scenario, the number of prevented cases could be as much as three quarters of the actual case count, and a reduction of 18-23% in burden on the healthcare system.

Economic analysis shows the mass testing pilot was cost-effective. This compares favourably with cost effectiveness thresholds such as the quoted National Institute for Health and Care Excellence (NICE’s) threshold of £30,000 per QALY gained, or the health production cost for the NHS in England, which is said to be around £15,000 per QALY gained.^29^ Even with pessimistic assumptions, the intervention is likely to be cost effective with a worst case scenario of £5,562 (£3,339 - £8,280 per QALY).

### Strengths and weaknesses of the study

This study is the first to evaluate the effectiveness of LFD mass testing by modelling the number of cases and healthcare burdens prevented and quantifying the benefits in terms of quality life years gained and healthcare costs saved. A strength of the present study, is that it uses real-world data from a mass testing pilot and regional estimates of Rt from actual case counts, as opposed to simulations. This eliminates many of the assumptions relating to the population and the costs and strategy of the mass testing programme. The main assumptions of our model were on the dynamics of onward transmission for which we validated with regional data where available.

A comparable economic analysis has not yet been conducted on the Liverpool pilot, however an interim analysis using Bayesian time-series modelling estimated a small non-significant reduction in cases and hospitalisations due to mass-testing.^2^ Our analysis shows that even a very small number of cases prevented can prove substantially cost-effective. Applying our model to the Liverpool data will therefore be prudent to determine if this is the case in Liverpool.

The model underlying these estimates makes a number of assumptions about the transmission dynamics of SARS-CoV-2. For example, the transmissibility of SARS_CoV-2 and the level of adherence to testing and self-isolation in symptomatic cases. Where possible, we used existing regional data to back up these assumptions. However, there is considerable uncertainty surrounding the definition of some of the parameters used. Our sensitivity analysis (See Appendix B) shows that the model is very sensitive to the relative infectability of asymptomatic cases, which is one parameter for which there is high uncertainty^17^ and is inconsistently defined.^30^ This makes the output of the model particular susceptible to errors in this parameter and should be considered when evaluating these estimates.

There are some important assumptions to consider in interpreting the results. Not all LFD positives had confirmatory PCR tests. Although we attempt to use PPV to estimate the proportion of additional true positives, this method could lead to overestimation of the number of cases prevented. The impact of contact tracing on behaviours of asymptotic cases was not considered. We assumed all asymptomatic cases would fail to isolate, but contact tracing would advise contacts of cases to self-isolate and be tested, so a proportion of asymptomatic cases would self-isolate without the mass testing intervention.

Another factor that was not considered in the model is differences in transmissibility between age and demographic groups. These groups will likely differ in their likelihood of transmitting and acquiring an infection. Such considerations would require generating Rt estimates for sub-populations for which sample sizes are likely to be too small. Also, the impact of new variants of concern and vaccine roll-out were not considered. These were not factors at the time of the pilot, but have subsequently become significant factors in the transmission dynamics of COVID-19. We have been quite conservative and not included costs of non-hospital cases or long COVID cases, and the QALY losses from long COVID cases are also quite conservative.

The true costs and QALYs lost from COVID-19 are likely to be higher than these estimates which do not include post-hospital rehabilitation, the likely full implications of multi-organ damage or informal care costs, productivity and wages lost. The indirect opportunity costs for people whose NHS treatment is delayed because of treating COVID-19 patients is not included. Overall, if more costs were included, the benefits would be likely to be greater. There are other onward costs of the COVID-19 system that are not included in this such as the costs of contact tracing, but these would fall both onto the intervention costs side, and on the cost savings side of any cost effectiveness equation, because preventing cases would reduce the need for future contact tracing. We did not include equity impacts in the calculation of costs and benefits; Merthyr Tydfil has a deprived population so adding an additional weighting for equity would likely mean that the benefits would be greater.^31^ However, it may also mean that people are exposed to more competing risks of morbidity and mortality. We have not included the benefits to individuals in Merthyr Tydfil from employment in supporting the mass testing programme. There is evidence from another combined modelling and economic analysis study mass testing has economic benefits by reducing the number of uninfected workers having to isolate.^32^ We did not carry out probabilistic sensitivity analysis or vary the economic parameters except in one-way sensitivity analyses; so the range of results is influenced only by the upper and lower estimates of the epidemiological modelling.

### Implications for policymakers and future research

The central estimate of net monetary benefit was £5.8 million which means a benefit-cost ratio of around 11 for the £516,000 that the programme cost, or a return on investment of around £10.30 per £1 spent. This is in line with the finding that public health interventions often have a high return on investment.^33^

These encouraging results come despite the poor sensitivity of the LFD test. We were unable to calculate sensitivity from the pilot data due to no systematic PCR follow-up for LFD negative results. However, in a community setting, sensitivity is likely to be highly variable and can be as low as 3% [@^7^. The implication of our contrasting results is that consideration of mass testing programmes should be based on other factors, in addition to test sensitivity.

While generalisability of these finding to other regions with different socio-demographic profiles and under different prevalence scenarios needs further investigation, it is clear that mass testing, in conjunction with other NPI measures, should be considered an important tool in control of COIVD19.

The cost-effectiveness of mass testing will likely be influenced by specific demographics and local healthcare policies. In particular, the school mass testing programme currently undertaken in the UK should be evaluated for cost-effectiveness. Of particular concern is the impact of disease prevalence. The Merthyr Tydfil pilot was conducted at a time of particularly highly prevalence (peak incidence during the pilot was 282 cases per 100,000 population). Had the pilot been conducted more recently, when prevalence was lower, the cost-effectiveness will likely be reduced. Supplementary simulation work suggest that the mass testing program would no longer be cost-effective if background prevalence had been lower than 2% (see Appendix C). In addition to increasing the number of false negatives, the number of false positives will increase in a low prevalence scenario. This may have further detrimental societal impact beyond cost-effectiveness (e.g. lost wages, productivity etc.). The confounding effects of vaccine roll-out and variants of concern should be considered. Since the majority of the benefits are through prevented deaths, if vaccines reduce the infection fatality ratio, the cost effectiveness of preventing transmission will decrease. Estimating cost-effectiveness in these different scenarios will be useful for deriving thresholds of community prevalence at which mass testing will become cost-effective, which will benefit local policy makers when planning future mess testing programmes.

Another aspect our analysis does not consider is the potential behavioural impact of a negative LFD test result. While a positive LFD test result will motivate an individual to self-isolate and reduce further transmission, a negative LFD test result may lead to a false sense of assurance and individuals engaging in more risky behaviour. Further research into the impact of such behavioural changes is therefore warranted.

## Conclusions

Despite the poor sensitivity of the LFD test, its use in mass testing appears to have helped prevent a number of secondary cases during the Merthyr Tydfil pilot, which occurred during a period of high infection prevalence. A non-negligible number of hospitalisations and deaths are also likely to have been prevented. Economic analysis demonstrates that the pilot was a highly cost-effective exercise. However, there are a number of parameters, in particular prevalence, which are likely to significantly influence cost-effectiveness. Future work to identify thresholds of cost-effectiveness across these parameters will provide policy makers valuable guidance in planning of future mass testing programmes.

## Data Availability

Aggregated data on Covid-19 cases, hospitalizations and deaths is available on the Public Health Wales Rapid Covid-19 Surveillance Dashboard.
Further anonymized data can be made available on reasonable request.

https://public.tableau.com/profile/public.health.wales.health.protection#!/vizhome/RapidCOVID-19virology-Public/Headlinesummary

## Declarations

## Acknowledgements

We thank all those who contributed to mass testing pilot including Merthyr Tydfil County Borough Council, Rhondda Cynon Taf County Borough Council, Cwm Taf Morgannwg University Health Board, Welsh Ambulance Service Trust, Public Health Wales, Welsh Government, Schools, Police, Third Sector and Military personnel. Additional thanks to Malorie Perry, Arthur Duncan-Jones, Craiger Solomons, James Anderson, Drew Turner and Laia Fina for assistance with data access and processing. Thanks to Jane Salmon and Christopher Williams for valuable comments on the manuscript.

## Authors’ contribution

MD contributed to the conception and implementation of the main data analysis and modelling and writing the manuscript. BC Contributed to conception and implementation of the economic analysis and writing the manuscript. AG contributed to conception and implementation of the pilot which provided the data for this study. KN contributed to conception and implementation of the pilot which provided the data for this study. DRhT contributed to conception of the overall study, project supervision and writing the manuscript.

## Data Availability

Aggregated data on Covid-19 cases, hospitalizations and deaths used in this study are available on the Public Health Wales Rapid Covid-19 Surveillance Dashboard. https://public.tableau.com/app/profile/public.health.wales.health.protection/viz/RapidCOVID-19virology-Public/Headlinesummary

Further anonymized data can be made available on reasonable request.

## Consent for publication

Not applicable

## Appendix A: Mathematical description of the model

The estimated number of estimated cases prevented on day *t*, denoted *x*_*prev*_(*t*), takes the form of an ordinary differential equation. The number of infections seeded on each day is computed differently for 1^st^ generation infections 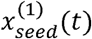 and higher generation infections 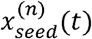.

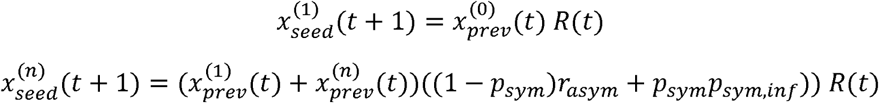

where *R*(*t*) is the reproduction number, *p*_*sym*_ is the proportion of cases that are symptomatic, and *r*_*asym*_ is the relative infectability of asymptomatic cases relative to symptomatic cases. The number of asymptomatic cases arising from prevented cases, given by *x*_*prev*_(*t*)(*1* − *p*_*sym*_) is assumed to contribute to the next generation of seeded cases. Additionally, a smaller number of symptomatic cases will also contribute to the next generation of seeded cases. This is given by *x*_*prev*_(*t*)(*p*_*sym*_*p*_*sym,inf*_), where *p*_*sym,inf*_ is the proportion of symptomatic cases that contribute to new cases. *p*_*sym*_ and *p*_*sym,inf*_ are assumed to be constant.

The initial number of infections projected (the 0th generation infections), 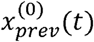) are given by the number of asymptomatic LFD tests with confirmatory positive PCR test result, *n*_*LFDtest*+*PCR*_(*t*), and with those no confirmatory PCR tests *n*_*LFDtest*_(*t*), the positivity prediction value *p*_*PPV*_, the proportion estimated to be asymptomatic, given by *1* − *p*_*sym*_, the proportion of symptomatic cases and still contributing to new infections *p*_*sym,inf*_ and the relative infectability of asymptomatic cases *r*_*sym*_.

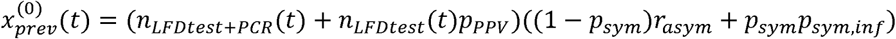

*x*_*prev*_(*t*) for other generations is computed from the convolution of *x*_*seed*_(*t*) with the log-normal model of the distribution of infections over time, *p*_*inf*_(*t*) *=* LognormalPDF(*t;µ,σ*^*2*^). For 1st generation infections, only the portion of the distribution from the median *e*^*µ*^ is included in the convolution.

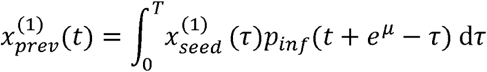

For nth generation infections, the whole distribution is included in the convolution.

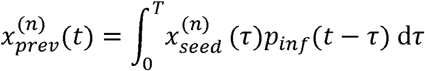

where *T* is the length of the interval being analysed.

The total number of prevented cases for each day is then obtained from the sum of prevented 1st and nth generation infections.

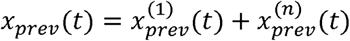

## Appendix B: Sensitivity analysis

Some of the assumptions underlying the analysis (see: Box 1) have a high degree of uncertainty. All these parameters depend on to categorisation of symptomatic and asymptomatic cases. There are nuances in the definitional boundary between symptomatic and asymptomatic infection across studies,^30^ including separate categorisations of presymptomatic and paucisymptomatic cases which were not made in this analysis. These nuances make this set of parameters particularly challenging to quantify. The parameters of concern are:

### 1. The proportion of asymptomatic cases that become symptomatic (assumption 8)

This value of 0.44 was obtained from the follow up case-control study undertaken at the same time as the mass testing.^16^ This estimate is chosen due to its highly specific to the geography and demography of the mass testing sample. However, this is quite a high estimate compared to other estimates in the literature. In the systematic review of 34, about three quarters of asymptomatic positive cases, never go on to develop symptoms. We therefore test against a lower best-case estimate of 0.25.

### 2. The proportion of symptomatic cases who contribute to further transmission (assumption 7)

This value of 0.2 was obtained from contact tracing data in the Merthyr Tydfil LA, cross referencing contacts and identifying which contacts originate from symptomatic positive cases. Again, this is the chosen estimate for the main analysis as it is highly specific to the geography and demography of the mass testing sample. However, incompleteness in contact reporting means this value is likely to be under-estimated. A cross-sectional survey in the UK by 35 indicate the proportion of symptomatic cases who fail to self-isolate is much higher (0.75). We therefore use this as a best-case alternative for this parameter.

### 3. The relative infectability of asymptomatic cases compared to symptomatic cases (assumption 2)

The value of 0.58 was obtained from the meta-analysis of 17. However, this estimate has very large confidence intervals (0.34 - 0.99, with the upper bound treating the two groups as virtually equal in infectability. We do not have robust way of estimating this parameter in the mass testing sample, so we use the central estimate from this meta-analysis as the main parameter and the bounds of the confidence interval from this meta-analysis as best-case and worst-case estimates.

Three parameters were highlighted for sensitivity analysis and varied to produce worst-case and best-case scenarios for the number of cases prevented and associated healthcare outcomes (worst case being highest number of cases prevented). Absolute and relative changes and corresponding sensitivities were computed for the best-case and worst-case scenarios for each parameter individually, with the base scenario being that used in the main analysis. The parameters used for the best- and worst-case scenarios used in the main analysis are shown in 7.

Results of sensitivity analysis are shown in Table 6. The first two parameters tested (proportion of asymptomatic cases that become symptomatic and proportion of symptomatic cases who contribute to further transmission) produced small relative sensitivity values (magnitude below 1). The number of cases prevented when adjusting these values to less conservative estimates are 424 and 644, respectively (representing increases of 18% and 79% of the original estimate). However, sensitivity to the relative infectability of asymptomatic cases was very large (greater than 2). The number of estimated cases prevented in the less conservative scenario is 1067 (an increase of 196%). The model is therefore particularly sensitive to this parameter and so uncertainly in its estimation should be carefully considered when evaluating estimates produced by this model.

**Table 6:** Results of sensitivity analysis.

| Parameter description | Original parameter | New parameter | Absolute parameter change | Relative Parameter change | New output | Absolute output change | Relative output change | Absolute sensitivity | Relative sensitivity |
| --- | --- | --- | --- | --- | --- | --- | --- | --- | --- |
| Proportion of asymptomatic cases who become symptomatic | 0.439 | 0.250 | -0.189 | -0.431 | 424 | 64 | 0.178 | -338 | -0.412 |
| Proportion of symptomatic cases who contribute to further transmission | 0.201 | 0.751 | 0.550 | 2.735 | 644 | 284 | 0.789 | 516 | 0.288 |
| Relative infectability of asymptomatic cases, compared to symptomatic cases | 0.580 | 0.990 | 0.410 | 0.707 | 1067 | 707 | 1.964 | 1724 | 2.778 |

**Table 7:**
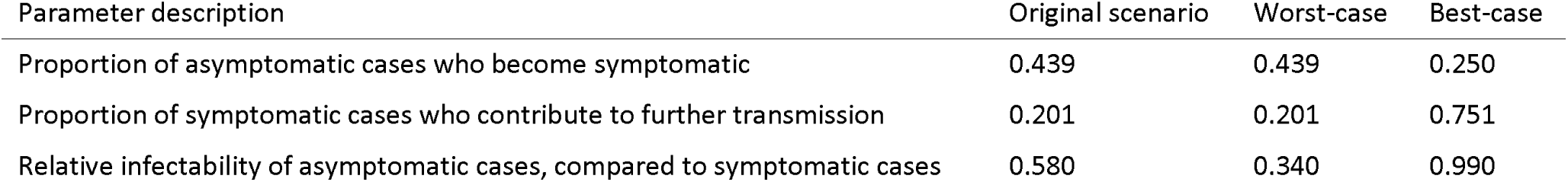
Parameters varied to produce best- and worst-case scenarios.

## Appendix C: Mapping Cost-effectiveness thresholds

In order to find approximate thresholds of cost-effectiveness, a set of simple simulations were carried out to across a space across four key parameters that affect the cost efficiency, in order to map cost-effectiveness across these parameters. LFD test results for each set of parameters were simulated. Prevalence, reproduction time (Rt), test sensitivity and test uptake. The number of asymptomatic individuals with a positive LTD test result is given by

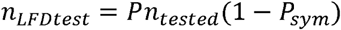

where *P* is the prevalence and *n*_*tested*_ is the number tested, which is given by

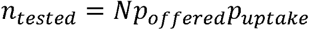

Where *N* is the total population, *p*_*offered*_ is the proportion offered tests and *p*_*uptake*_ is the proportion of uptake. *p*_*offered*_ *= 0*.*9*. We assume that no confirmatory PCR tests are performed, and that number of true LFD positives is as given by the PPV value *p*_*PPV*_, which was computed for the given test sensitivity and specificity values. Test specificity was fixed at 99%.

The simulated values of *n*_*LFDtest*_ were uniformly distributed over a *T= 30* day period, so that *n*_*LFDtest*_(*t*) *= n*_*LFDtest*_*/T*. The simulated *n*_*LFDtest*_(*t*) and *p*_*PPV*_ values were then fed into the model as described in appendix A. All other model parameters were as used in the original analysis. The same time-lagged model used in the main analysis was used to estimate hospitalisations and deaths prevented. The same operational costs, costs per test and QALYs from the main analysis was used to estimate ICER values.

The parameters ranges simulated were: Prevalence: [1%,2%,3%,4%,5%], Rt: [0.5,1.0,1.5,2.0,2.5], test sensitivity: [5%, 25%, 50%] and test uptake: [25%, 50%, 75%]. The simulated ICERs were mapped across the four parameters (Figure 4). ICERs were grouped into four levels of cost-efficiency: very high (<£0), high (£0-£15k), marginal (£15k-£30k) and none (>£30k). The approximate position of the mass testing pilot (Rt ≈ 1, prevalence ≈ 0.03, test sensitivity ≈ 50%, test uptake ≈ 50%) is indicated.

**Figure 4:**
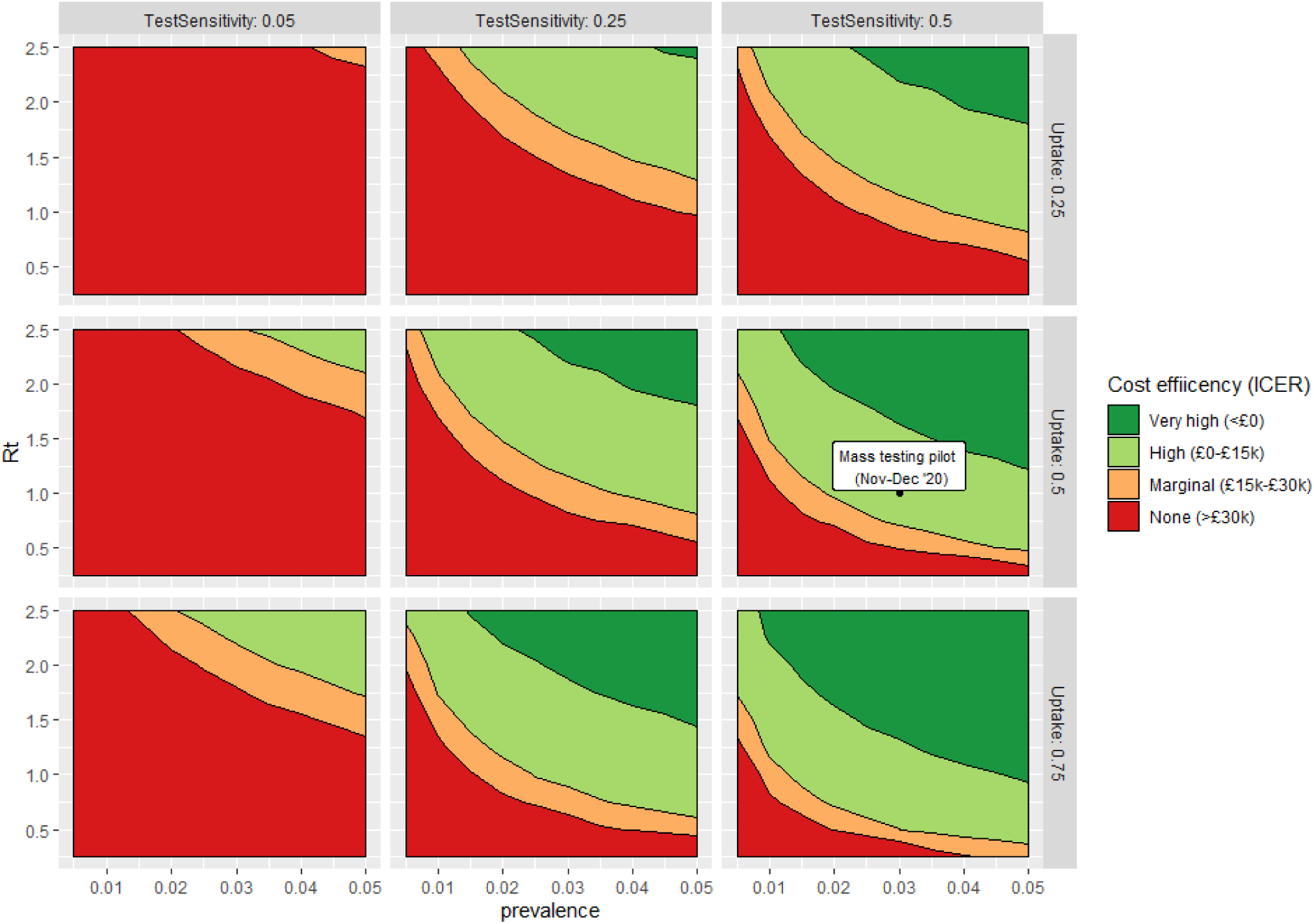
**Simulated ICER levels across four parameters: Rt, prevalence, test sensitivity and test uptake. All other model parameters fixed as defined in the main analysis. The approximate position of the mass testing pilot in the parameter space is indicated**.

The overall trend of cost effectiveness increases as prevalence and Rt increases. Cost-effectiveness shows a very high effect of test sensitivity, with a sensitivity of 5% making most of the parameter space non-cost effective. Test uptake has more impact when test sensitivity low. At 50% sensitivity, there is only a small effect of test update.

With respect the conditions around the mass testing pilot, cost-effectiveness is predicted to be high or very high (ICER < £15k) when prevalence is above 2%, with an Rt of 1, or when Rt is above 0.75, with a prevalence of 3%.

